# Brain Structural Underpinnings of the Differential Effects of Threat and Deprivation on Executive Function: A UK Biobank Study

**DOI:** 10.1101/2025.09.03.25333003

**Authors:** Hao Qin, Jiaying Xu, Jiesi Wang, Weiwen Wang, Hang Xu

## Abstract

Childhood adversity is known to impair Executive Function (EF) and brain development, but the distinct effects of deprivation versus threat remain unclear. In this study, we used data from the UK Biobank (UKB) to examine how these two adversity subtypes relate to performance across multiple EF domains: working memory, cognitive flexibility, problem solving, rule comprehension, and to regional brain volumes. We found that greater deprivation was linked to poorer performance across all EF domains and to structural changes marked by enlarged striatal volumes and reduced left dentate gyrus (DG) volume. Notably, left DG volume marginally mediated the relationship between deprivation and General Executive Function (GEF). In contrast, higher threat exposure was linked only to better verbal working memory and GEF, as well as reduced volumes in multiple thalamic nuclei. right ventromedial (VM) volume significantly mediated the association between threat exposure and GEF. These results indicate that deprivation exerts broad negative effects on EF, whereas threat produces more selective impacts, potentially reflecting adaptive enhancement of specific functions, DG and VM serve as distinct neuroanatomical mediators for these two forms of childhood adversity.

## Introduction

Executive Function (EF) comprises higher-order cognitive processes such as working memory, cognitive flexibility, and inhibitory control, all of which support goal-directed behavior and adaptation to changing environments (Miyake et al., 2000). Notably, structural and functional plasticity of the brain network involved in cognitive control undergoes rapid and profound development during childhood and adolescence, making it a sensitive time window for adverse life events to influence EF development (Ferguson et al., 2021). Childhood adversity, broadly defined as early-life experiences that disrupt healthy development—including various forms of neglect and abuse—has been consistently linked to long-lasting EF impairments, altering its developmental trajectory from childhood into adulthood (Pollak et al., 2000). Most studies quantify adversity using a cumulative risk approach. Although higher scores predict greater EF impairment, this approach assumes all experiences have equivalent effects, ignoring the distinct neurodevelopmental pathways of different adversity types (Lund et al., 2020). For example, three instances of physical abuse and three of emotional neglect yield the same score yet affect brain development in fundamentally different ways (Shin et al., 2018).

Recognizing this limitation has led to dimensional frameworks such as the Dimensional Model of Adversity and Psychopathology (DMAP), which distinguishes adversity types based on their underlying mechanisms (McLaughlin et al., 2014; Sheridan & McLaughlin, 2014). DMAP classifies adversity into two core dimensions: Deprivation, defined as the absence of expected environmental inputs (e.g., physical or emotional neglect), Threat, defined as experiences involving harm or threat to physical or emotional safety (e.g., physical, emotional, or sexual abuse). Accumulating evidence suggests these dimensions have dissociable effects on EF—deprivation is more consistently associated with severe and broad EF deficits, whereas the impact of threat is weaker, inconsistent, and in some cases even protective (Dannehl et al., 2017; Feeney et al., 2013; Hawkins et al., 2021; Johnson et al., 2021; Lin et al., 2022; Vogel et al., 2021). Yet, few studies have measured deprivation and threat within the same framework, leaving open critical questions about their differential impacts and underlying mechanisms.

The DMAP, informed by animal research, initially hypothesized that threat would be associated with volumetric differences in subcortical regions such as the amygdala and hippocampus, whereas deprivation would be linked to increased cortical thickness in association cortices (McLaughlin et al., 2014). However, empirical findings have not fully supported this pattern. For instance, in a large pediatric sample, Machlin found no association between threat and amygdala or hippocampal volume, while deprivation was related to increased cortical thickness in the occipital cortex, insula, and cingulate cortex (Machlin et al., 2023), Other studies report that deprivation can also affect subcortical structures, including volumetric reductions in the striatum and hippocampus (Mowery et al., 2017; Staff et al., 2012), consistent with the hippocampus’s known sensitivity to environmental stimulation and stress hormone exposure during development. For threat, some evidence implicates alterations in subcortical relay structures such as the thalamus, a hub for sensory integration and salience processing (Zhu et al., 2018). Many of these regions—including the hippocampus, striatum, and thalamus—are also engaged during core cognitive operations, including executive control (Nelson, 2021). Despite these insights, existing evidence remains fragmented across samples, age groups, and analytic approaches. The specific structural pathways through which deprivation and threat shape EF performance remain insufficiently understood, underscoring the need for studies that assess multiple adversity dimensions within the same cohort and directly test their distinct neural substrates.

Large-scale, multimodal datasets are critical for disentangling these controversial issues. The UK Biobank (UKB; see https://biobank.ndph.ox.ac.uk) provides an unparalleled platform, combining retrospective reports of childhood experiences that differentiate deprivation from threat with comprehensive EF assessments and high-quality neuroimaging. With over 500,000 participants aged 40–69 years at baseline, the UKB offers a sample size large enough to detect subtle brain – behavior associations with high statistical power. Its neuroimaging data — collected using standardized protocols across multiple modalities (e.g., structural MRI, diffusion MRI) — enables precise mapping of cortical and subcortical changes linked to different adversity dimensions. Moreover, the inclusion of detailed health, lifestyle, and genetic data allows for rigorous control of potential confounders.

In the present study, we leverage these strengths to systematically compare how deprivation and threat are associated with EF performance and to identify the neural substrates that mediate these relationships, thereby advancing our understanding of the distinct neurodevelopmental pathways shaped by early-life adversity.

## Methods

### Participants

Data for this study were obtained from the UKB, a large−scale biomedical resource comprising health and lifestyle information. Available variables include demographic characteristics (e.g., sex, age, ethnicity), medical history and disease status, cognitive function assessments, and neuroimaging measures. The UK Biobank Research Ethics Committee and the Cambridge University Human Biology Research Ethics Committee granted ethical approval for use of these data (see UK Biobank Ethics, n.d.). All participants provided written informed consent prior to enrollment.

We initially downloaded data for 502,235 participants from the UKB. To avoid confounding effects of cardiovascular disease, diabetes mellitus, and cancer on brain structure, we excluded anyone who self−reported a doctor−diagnosed cardiovascular condition (Data Field:6150), diabetes mellitus (Data Field:2443), or cancer (Data Field:2453), yielding a healthy−control sample of N = 316,369. For behavioral analyses, we included participants from this subsample who completed the childhood adversity assessment and at least one EF test during their imaging visit. For neuroimaging analyses, we included only those participants with usable T1−weighted structural magnetic resonance imaging (MRI) scans. Because completion rates for EF testing and MRI acquisition varied, sample sizes differed across analyses; exact Ns for each test are reported in the Results section.

### Childhood Adversity Assessment

Childhood adversity was assessed in the UKB using the Childhood Trauma Screener−5 items (CTS−5). Items were rated on a five-point Likert scale; participants with any missing response were excluded. The CTS-5 includes five items: Emotional abuse (“Felt hated by a family member as a child”; Field:20487), Physical abuse (“Physically abused by family as a child”; Field:20488), Sexual abuse (“Sexually molested as a child”; Field:20490), Emotional neglect (“Felt loved as a child”; Field:20489; reverse-scored), Physical neglect (“Had someone to take them to the doctor when needed as a child”; Field:20491; reverse-scored). For participants with complete CTS−5 data, we computed three summary scores: cumulative adversity (sum of all five items), threat exposure (sum of the three abuse items), and deprivation exposure (sum of the two neglect items). Summary scores were coded in a positive direction, such that higher values indicate more severe childhood adversity.

### Executive Function Assessment

EF was assessed with six tasks administered during the MRI session to align behavioral measures with imaging acquisition. The tasks and their targeted domains were: N-back Test (digit working memory); Pairs Matching Test (visuospatial working memory); Paired Associates Learning Test (verbal working memory); Trail Making Test (cognitive flexibility); Tower Rearranging Test (problem solving); and Matrix Test (rule understanding). Detailed protocols and primary outcome measures are provided in the Supplementary Material S1 Appendix 1.

### Magnetic Resonance Imaging Assessment

Magnetic resonance imaging (MRI) data were acquired on a single 3-T Siemens Skyra scanner (Erlangen, Germany). T1-weighted images were collected using a magnetization-prepared rapid gradient-echo (MPRAGE) sequence with 1 × 1 × 1 mm isotropic voxels and a 208 × 256 × 256 acquisition matrix (Alfaro-Almagro et al., 2018).

Volumetric segmentation was performed using three complementary methods: FIRST (FMRIB’s Integrated Registration and Segmentation Tool): volumetric measures for all 14 subcortical regions (Category 1102); ASEG (FreeSurfer’s Automated Segmentation): retained 16 bilateral regions after excluding ventricles, cerebellum, brainstem, optic chiasm, corpus callosum, choroid plexus, vascular structures, and global measures (Category 190); and subcortical subfield segmentation (Freesurfer): volumes for thalamic nuclei, amygdala subunits, and hippocampal subfields (Category 191). Full details of regional brain volumes can be found in the Supplementary Material S1 Appendix 2.

To control for head size, we included total gray-plus-white matter volume (Data-Field 25010; FSL processing) as a covariate in FIRST analyses and total intracranial volume (Data-Field 26521; FreeSurfer) in ASEG and subfield analyses. All segmentation protocols and quality-control procedures follow the UKB imaging pipeline.

### Other Variables

Additional variables included self-reported demographic information (e.g., sex, age, body mass index [BMI], ethnicity); educational deprivation, measured using the England-specific Townsend education deprivation index; income deprivation, measured using the England-specific Townsend income deprivation index; and fluid intelligence, assessed via a standardized 13-item test (score range = 0-13). Current research suggests that while declines in fluid intelligence partially account for EF impairments, they do not fully explain all EF deficits. Although including fluid intelligence as a covariate may reduce the observed effects of EF impairments (Roca et al., 2013), this approach is essential for identifying the specific components of executive dysfunction that are independent of general cognitive ability (Roca et al., 2014).

### Statistical Analysis

We conducted all analyses in RStudio (R version 4.3.3). First, to derive a General Executive Function (GEF) factor, we performed principal-component analysis (PCA) on the six EF measures. To align scoring directions, visuospatial working memory and cognitive flexibility scores were multiplied by −1 prior to PCA (Friedman & Miyake, 2017). Before extraction, we evaluated the data’s suitability for PCA using the Kaiser–Meyer–Olkin (KMO) measure of sampling adequacy and Bartlett’s test of sphericity. Acceptable PCA conditions were defined as KMO values above .60 and a significant Bartlett’s test (p < .05), indicating sufficient shared variance among variables (ref). We retained components with eigenvalues greater than one and used the first principal component as a composite GEF score in subsequent analyses.

Next, we ran multiple linear regression models to test the differential effects of deprivation and threat on both EF and regional brain volumes. All models included age; sex; ethnicity; BMI; educational deprivation, index income deprivation index, and fluid intelligence as covariates. When testing deprivation effects, we additionally adjusted for threat severity; when testing threat effects, we adjusted for deprivation severity, minimizing confounding. Head-size covariates—total gray plus white matter volume (Data-Field 25010) for FIRST and total intracranial volume (Data-Field 26521) for ASEG and subfield analyses—were included in all imaging models.

Based on regression and partial-correlation results, we specified structural equation models (SEMs) in which EF was regressed on a candidate childhood adversity (X), a candidate mediator (M), and baseline covariates (age; sex; ethnicity; BMI; educational deprivation index; income deprivation index, and fluid intelligence). Head-size covariates were included as above. To isolate EF-specific effects, threat models adjusted for deprivation severity, deprivation models adjusted for threat severity.

Using bias-corrected bootstrap resampling (1,000 iterations), we estimated standardized direct, indirect, and total effects with 95% confidence intervals. Model fit was evaluated with chi-square-to-degrees-of-freedom ratio χ²/df; Comparative Fit Index (CFI); Tucker–Lewis Index (TLI); Root Mean Square Error of Approximation (RMSEA); and Standardized Root Mean Square Residual (SRMR). Criteria for an excellent fit were: χ²/df < 2; CFI and TLI > .95; RMSEA and SRMR < .05. Criteria for an acceptable fit were: χ²/df < 3; CFI and TLI > .90; and RMSEA and SRMR < .08. Given the sensitivity of χ²/df to large samples (Klumparendt et al., 2019), models meeting at least two other criteria were also deemed acceptable. To control the false discovery rate (FDR) in our volumetric analyses, p values were adjusted using the Benjamini–Hochberg procedure. Continuous variables are presented as mean ± standard deviation; categorical variables as counts (percentages).

## Results

### Baseline Characteristics of Participants

Table 1 presents the demographic information of the 316,369 participants. Of these, 180,068 (56.9%) were female, with ages ranged from 38 to 73 years (M = 54.89, SD = 7.45). The sample was predominantly White (N = 287,297; 90.8%). In addition to demographics, the table also summarizes key study variables, including dependent variables (performance on various EF tasks), the main independent variable (childhood adversity measured by CTS-5), and covariates (educational deprivation index, income deprivation index, BMI, TIV, and fluid intelligence scores).

**Table 1.**
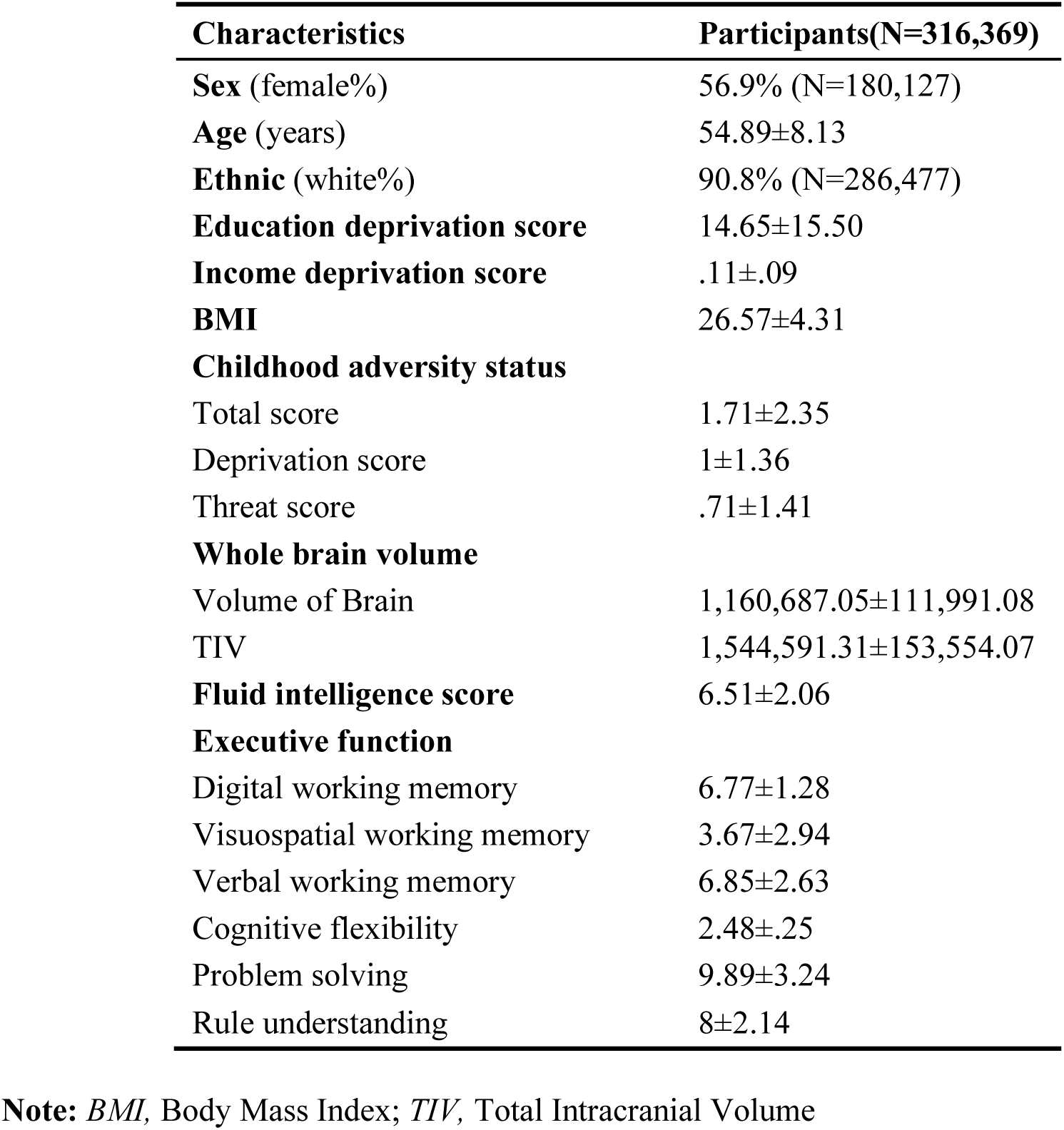
Demographic Characteristics.

| **Characteristics** | **Participants(N=316,369)** |
| --- | --- |
| **Sex** (female%) | 56.9% (N=180,127) |
| **Age** (years) | 54.89±8.13 |
| **Ethnic** (white%) | 90.8% (N=286,477) |
| **Education deprivation score** | 14.65±15.50 |
| **Income deprivation score** | .11±.09 |
| **BMI** | 26.57±4.31 |
| **Childhood adversity status** |  |
| Total score | 1.71±2.35 |
| Deprivation score | 1±1.36 |
| Threat score | .71±1.41 |
| **Whole brain volume** |  |
| Volume of Brain | 1,160,687.05±111,991.08 |
| TIV | 1,544,591.31±153,554.07 |
| **Fluid intelligence score** | 6.51±2.06 |
| **Executive function** |  |
| Digital working memory | 6.77±1.28 |
| Visuospatial working memory | 3.67±2.94 |
| Verbal working memory | 6.85±2.63 |
| Cognitive flexibility | 2.48±.25 |
| Problem solving | 9.89±3.24 |
| Rule understanding | 8±2.14 |

**Note:** *BMI*, Body Mass Index; *TIV*, Total Intracranial Volume

### Calculation of General Executive Function Score

Figure 1 displays the results of PCA conducted on the six EF task scores. The data were deemed suitable for PCA, as indicated by a KMO measure of sampling adequacy of .773 and a significant Bartlett’s test of sphericity, χ²(15) = 17,665.78, p < .001. One component with an eigenvalue greater than 1 was extracted, accounting for 36.3% of the total variance. This component was labeled GEF and was used as a composite EF score in subsequent analyses.

**Figure 1.**
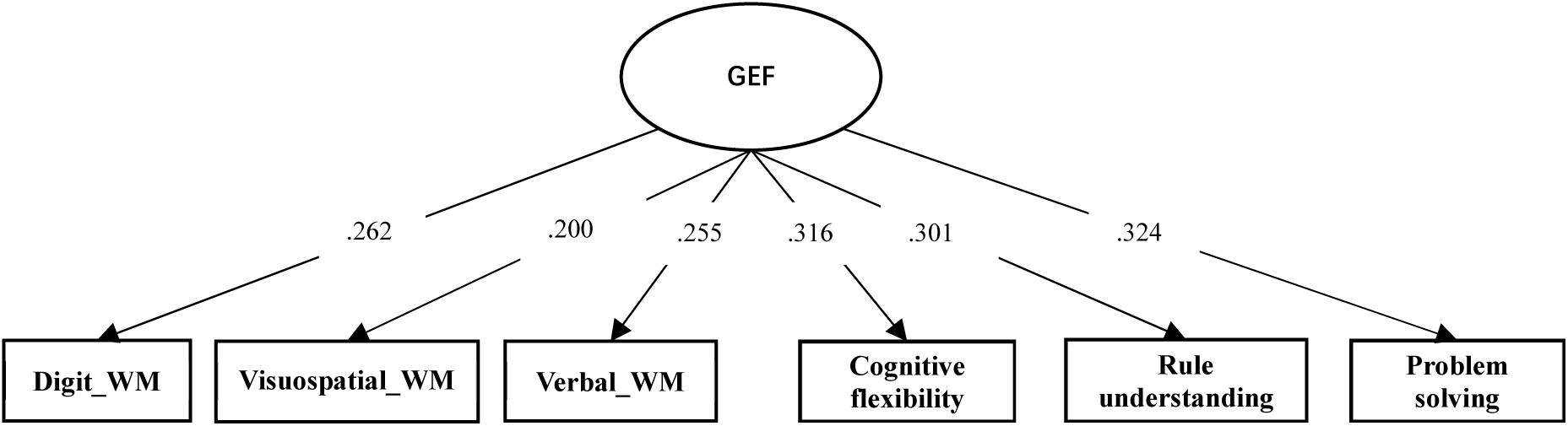
Principal Component Analysis of GEF. **Note:***WM*,Working Memory; The number on the arrow represents the factor loadings

**Figure 2.**
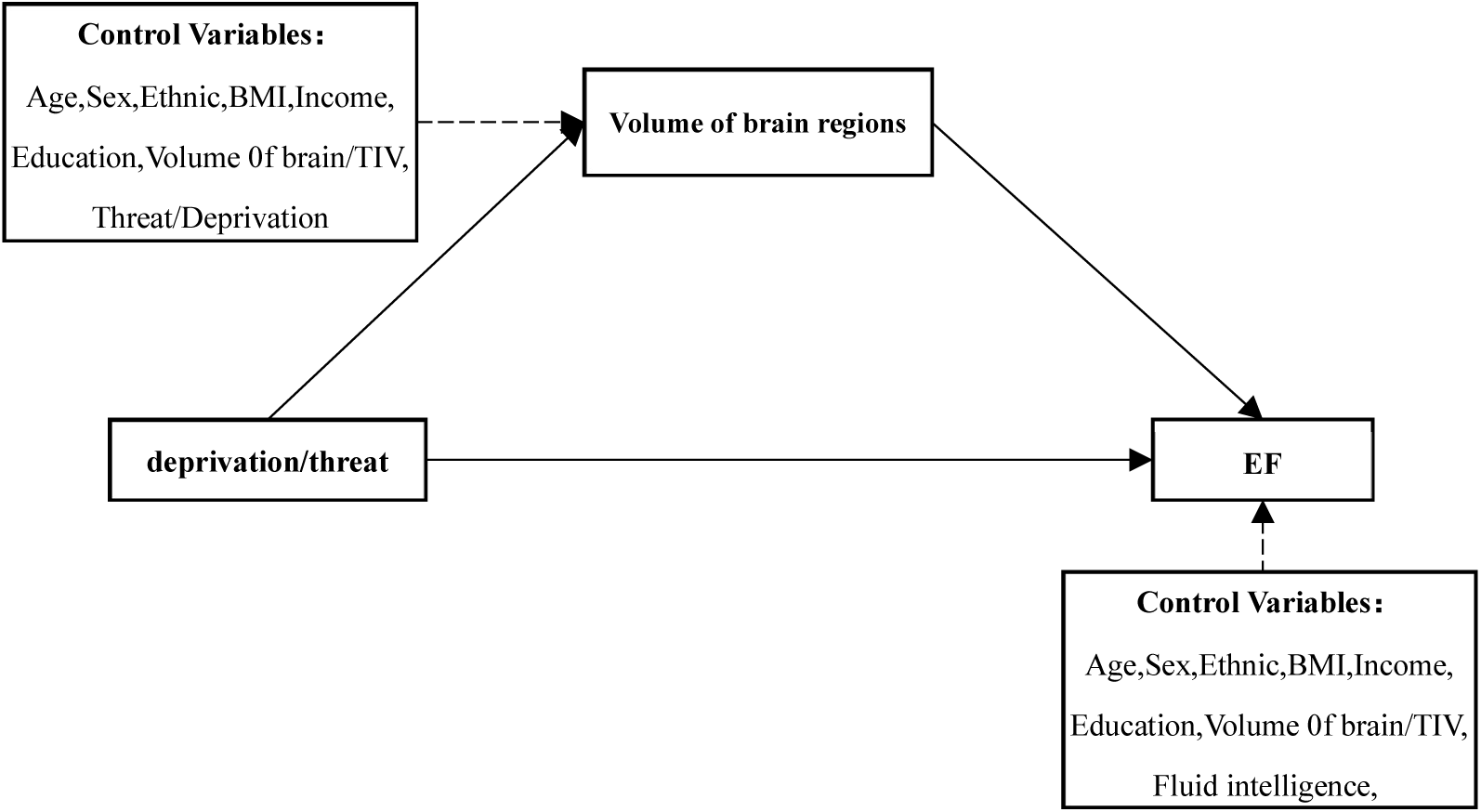
Mediation Analysis Model setting.

### The Effects of Deprivation/threat on Executive Function

Higher deprivation was significantly associated with poorer performance across all EF components: lower maximum digit span (β = *–.028*, 95% CI [*–.043, –.013*], *p* < *.001*); more errors in visuospatial working memory (β = *.037*, 95% CI [*.009, .065*], *p* = *.009*); fewer correct responses in verbal working memory (β = *–.090*, 95% CI [*–.116, –.063*], *p* < *.001*); longer completion time on the Trail Making Test (β = *.008*, 95% CI [*.005, .010*], *p* < *.001*); fewer correctly solved Tower Rearranging trials (β = *–.099*, 95% CI [*–.137, –.060*], *p* < *.001*) and fewer correct Matrix Test items (β = *–.060*, 95% CI [*–.081, –.039*], *p* < *.001*). Deprivation was also linked to lower scores on the GEF factor (β = *–.066*, 95% CI [*–.081, –.051*], *p* < .*001*). In contrast, threat exposure was positively associated only with verbal working memory (β = *.028*, 95% CI [*.003, .053*], *p* = *.030*) and the GEF factor (β = *.020*, 95% CI [*.006, .035*], *p* = *.005*), with no significant associations observed for the other EF measures (Full results are presented in Table 2).

**Table 2.** The effects of deprivation/threat on executive function.

| Dependent_variable | Deprivation |  |  | Threat |  |  |
| --- | --- | --- | --- | --- | --- | --- |
| | $\beta$ | 95%CI | Adj_R2 | $\beta$ | 95%CI | Adj_R2 |
| Digital working memory (N=17107) | -.028*** | [-.043, -.013] | .155 | .003 | [-.012, .017] | .155 |
| Visuospatial working memory (N=28368) | .037** | [.009,.065] | .048 | .001 | [-.026, .027] | .048 |
| Verbal working memory (N=22110) | -.090*** | [-.116, -.063] | .153 | .028* | [.003, .053] | .153 |
| Cognitive flexibility (N=21503) | .008*** | [.005,.010] | .204 | -.002 | [-.005, .000] | .204 |
| Problem solving (N=16542) | -.099*** | [-.137, -.060] | .150 | .036 | [-.001,.074] | .150 |
| Rule understanding (N=21988) | -.060*** | [-.081, -.039] | .223 | .017 | [-.003,.037] | .223 |
| GEF (N=16003) | -.066*** | [-.081, -.051] | .376 | .020** | [.006,.035] | .376 |

**Note:** \* $p < .05$ , \*\* $p < .01$ , \*\*\* $p < .001$ ; 95%CI, 95% confidence interval; Adj\_R<sup>2</sup>, adjusted R<sup>2</sup>

### The Effects of Deprivation/Threat on Brain Region Volumes

Table 3 shows that deprivation was associated with volumetric increases in FIRST-segmented striatal subregions, including the left caudate (β = *6.04*, 95% CI [*2.07, 10.02*], adjusted *p* < *.05*), right caudate (β = *7.27*, 95% CI [*3.08, 11.47*], adjusted *p < .01*), and left putamen (β = *7.37*, 95% CI [*2.20, 12.54*], adjusted *p < .05*), with a marginal increase in the right putamen (β = *5.83*, 95% CI [*2.20, 12.54*], adjusted *p = .075*). Conversely, deprivation predicted reductions in hippocampal subfield volumes based on FreeSurfer subsegmentation, including the left Granule Cell-Molecular Layer-Dentate Gyrus body (GC-ML-DG body) (β = *–.28*, 95% CI [*–.49, –.08*], adjusted *p = .075*), right GC-ML-DG body (β = *–.30*, 95% CI [*–.51, –.10*], adjusted *p = .069*), and the left subiculum body (β = *–.62*, 95% CI [*–.98, –.26*], adjusted *p < .05*).

**Table 3.**
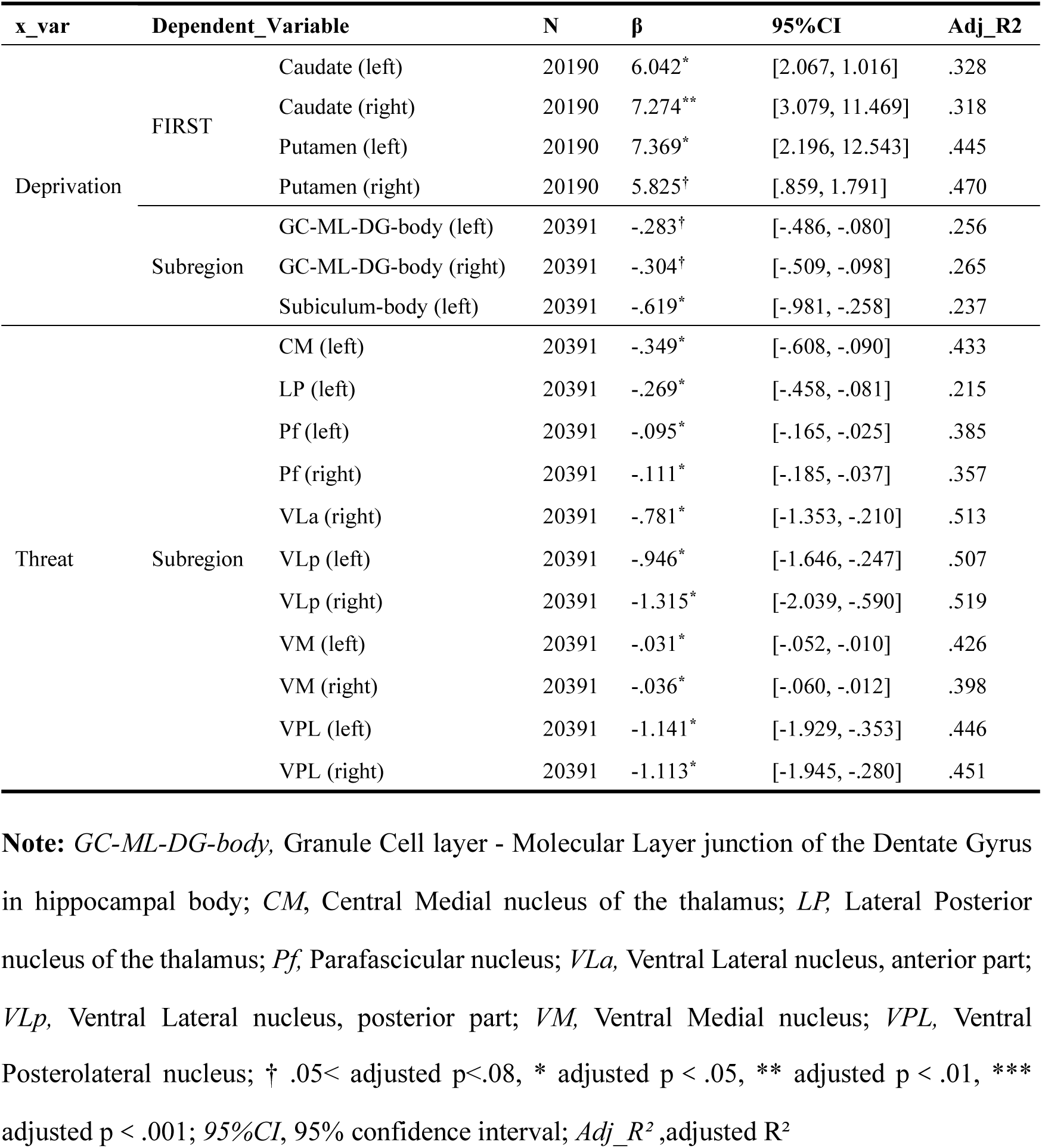
The effects of deprivation/threat on brain region volumes.

**Tabel 4.** Total Effect, Direct Effect and Indirect Effect of deprivation on EF through brain region volumes.

| Predictor | Dependent variable | Mediator |  | Total effect | Direct effect | Indirect effect | Indirect 95%CI |
| --- | --- | --- | --- | --- | --- | --- | --- |
| **Deprivation** | Digital working memory | FIRST | Putamen (left) | -.0273** | -.0279*** | .0007* | [.0002, .0016] |
|  |  | Subregions | GC-ML-DG-body (left) | -.0315*** | -.0305*** | -.0010* | [-.0020, -.0002] |
|  |  |  | Subiculum-body (left) | -.0315*** | -.0309*** | -.0006* | [-.0014, -.0001] |
|  | Visuospatial working memory | FIRST | Caudate (right) | .0194* | .0189* | .0005* | [.0002, .0012] |
|  | Rule understanding | Subregions | GC-ML-DG-body (left) | -.0327*** | -.0322*** | -.0006† | [-.0014, -.0001] |
|  | GEF | Subregions | GC-ML-DG-body (left) | -.0521*** | -.0516*** | -.0005† | [-.0014, -.0001] |

**Note:** *GC-ML-DG-body*, Granule Cell layer - Molecular Layer junction of the Dentate Gyrus in hippocampal body; *CM*, Central Medial nucleus of the thalamus; *VM*, Ventral Medial nucleus; *VPL*, Ventral Posterolateral nucleus; *VLa*, Ventral Lateral Anterior nucleus; *VLp*, Ventral Lateral Posterior nucleus; *95%CI*, 95% confidence interval; † .05 < adjusted p < .08, \*p < .05, \*\*p < .01, \*\*\*p < .001

**Tabel 5.**
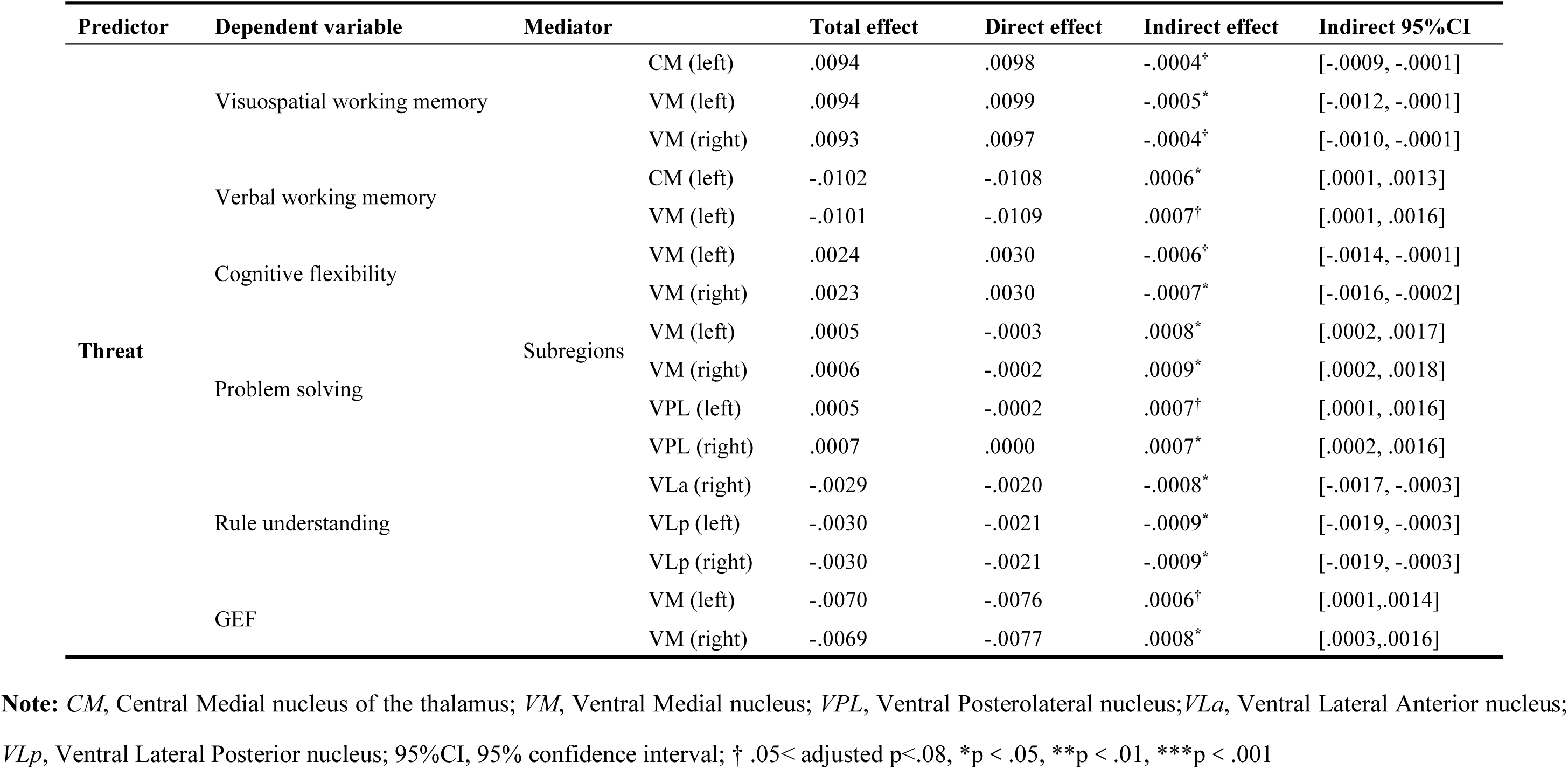
Total Effect, Direct Effect and Indirect Effect of threat on EF through brain region volumes.

Threat exposure, in contrast, was associated with decreased volumes across multiple thalamic nuclei in FreeSurfer subsegmentation, including the left centromedian nucleus (CM) (β = *–.35*, 95% CI [*–.61, –.09*], adjusted *p < .05*), left lateral posterior nucleus (LP) (β = *–.27*, 95% CI [*–.46, –.08*], adjusted *p < .05*), left parafascicular nucleus (Pf) (β = *–.10*, 95% CI [*–.17, –.03*], adjusted *p < .05*), right Pf (β = *–.11*, 95% CI [*–.19, –.04*], adjusted *p < .05*), right ventral lateral anterior nucleus (VLa) (β = *–.78*, 95% CI [*–1.35, –.21*], adjusted *p < .05*), left ventral lateral posterior nucleus (VLp) (β = *–.95*, 95% CI [*–1.65, –.25*], adjusted *p < .05*), right VLp (β = *– 1.32*, 95% CI [ *– 2.04, –.59*], adjusted *p < .05*), left ventromedial nucleus (VM) ( β = *–.03*, 95% CI [ *–.05, –.01*], adjusted *p < .05*), right VM ( β = *–.04*, 95% CI [ *–.06, –.01*], adjusted *p < .05*), left ventral posterolateral nucleus (VPL) (β = *–1.14*, 95% CI [*–1.93, –.35*], adjusted *p < .05*), and right VPL (β = *–1.11*, 95% CI [*–1.95, –.28*], adjusted *p < .05*).

### Mediation of Deprivation and Threat Effects on Executive Function via Brain Structures

Partial-correlation analyses (see Supplementary Materials S2) identified several regional brain volumes associated with EF, which were subsequently tested as mediators in structural equation models. Mediation analyses showed that both deprivation and threat were associated with EF through small but significant indirect effects involving distinct brain regions.

For deprivation, significant mediation effects were observed through both hippocampal subfields (the left GC-ML-DG-body and the left subiculum-body) and striatum structures (the left putamen and the right caudate). The left GC-ML-DG-body mediated the associations between deprivation and multiple EF components, including digit working memory (ab = *–.001*, *p < .05*), rule understanding (ab = −*.0006*, *p = .066*), and GEF (ab = −*.0005*, *p = .065*). The left subiculum-body mediated the associations between deprivation and digit working memory (ab = −*.0006*, *p < .05*). The left putamen mediated the association between deprivation and digital working memory (ab = *.0007*, *p < .05*), while the right caudate was mediated the association between deprivation and visuospatial working memory (ab = *.0005*, *p < .05*), indicating task-specific involvement of striatal subregions (see Table 2). All deprivation models demonstrated excellent fit (CFI > .90, TLI > .90, RMSEA < .05, χ²/df < 3).

For threat, significant indirect effects were primarily observed through thalamic subregions, including the VM, CM, VPL, and VL. The VM nuclei, both left and right, mediated the associations between threat and multiple EF components, including visuospatial working memory (left: ab = −.0005, *p* < .05; right: ab = −.0004, *p* = .064), verbal working memory (left: ab = .0007, *p* = .053), cognitive flexibility (left: ab = −.0006, *p* = .072; right: ab = −.0007, *p* < .05), problem solving (left: ab = .0009, *p* < .05; right: ab = .0009, *p* < .05), and GEF (left: ab = .0006, *p* = .063; right: ab = .0008, *p* < .05). The other nuclei showed more selective effects: the left CM mediating the associations with visuospatial working memory (ab = −.0004, *p* < .05) and verbal working memory (ab = .0006, *p* < .05); the VPL bilaterally mediated problem solving (left and right: ab = −.0004, *p* < .05); and within the VL, the VLp bilaterally mediated rule understanding (left: ab = .0007, *p* = .055; right: ab = .0007, *p* < .05), while the right VLa also mediated this effect (ab = −.0008, *p* < .05) (see Table 2). All threat models met CFI > .90, TLI > .90, and RMSEA < .05.

## Discussion

This study leveraged UKB data to systematically examine how childhood deprivation and threat differentially affect EF and its neural substrates. We found that deprivation significantly impaired all EF domains, whereas threat selectively influenced verbal working memory and GEF. These findings highlight marked heterogeneity in the cognitive consequences of adversity, suggesting that deprivation exerts broad negative effects while threat may produce more domain-specific or even context-dependent adaptive influences. Neuroanatomically, deprivation-related EF deficits were mediated by hippocampal and striatal alterations, while threat-related effects were primarily mediated through high-order thalamic nuclei. Together, these findings underscore the importance of considering deprivation and threat as distinct developmental exposures that shape EF via separable neural pathways.

Our results align with prior evidence showing broad and persistent EF deficits following deprivation (Machlin et al., 2019) but more variable effects of threat (Lin et al., 2022). Some studies even suggest that threat can facilitate specific EF domains, such as working memory (Dannehl et al., 2017; Feeney et al., 2013). Consistent with the “hidden talents” hypothesis (Ellis et al., 2022), our findings indicate that threat exposure may sharpen attentional and control processes under certain conditions. These results therefore support a more nuanced interpretation of adversity, incorporating adaptive as well as deficit-focused perspectives.

By examining subregional volumes within EF-related circuits, we identified distinct neurostructural mediators. Deprivation primarily affected EF through alterations in the dentate gyrus (DG) and striatal subregions. The DG is implicated not only in memory encoding and spatial processing but also in cognitive flexibility through its role in rapid memory updating and switching (Anacker & Hen, 2017; Hainmueller & Bartos, 2020). Experimental evidence further shows that DG is highly stress-sensitive, with rodent studies documenting ventral DG damage under stress (Tanti et al., 2013) and antidepressant treatments restoring function via neurogenesis (Boldrini et al., 2009). These findings reinforce the DG as an environmentally sensitive substrate underlying deprivation-related EF impairment. In contrast, threat effects were mediated through higher-order thalamic nuclei, particularly VM and CM regions, which are central hubs in thalamo-cortical loops supporting attention, arousal, and working memory (Sherman, 2005; Shine et al., 2023). Volume reductions in these nuclei may reflect remodeling of attention-related circuits in response to threat, with selective improvements in EF suggesting potential adaptive changes in threatening environments.

Unlike prior reports of deprivation-related prefrontal alterations (Machlin et al., 2023; McLaughlin et al., 2019), we did not observe significant mediation via prefrontal regions. This may reflect age-related normalization of early structural changes (Shaw et al., 2013), the limited sensitivity of volumetric indices compared with functional or microstructural measures, or indirect prefrontal involvement via subcortical nodes (e.g., dentate gyrus, thalamic nuclei) that are densely interconnected with prefrontal circuits (Rolls, 2022; Rolls et al., 2022). Thus, while the prefrontal cortex remains central to EF, our results suggest that in adulthood deprivation effects are more prominently expressed through hippocampal and thalamostriatal pathways than persistent prefrontal volume loss.

Although both deprivation and threat significantly influenced EF, the effect sizes were small and the mediation effects modest. Such subtle coefficients may nonetheless represent meaningful pathways, as EF is shaped by a wide range of genetic and environmental influences, with single adversity dimensions accounting for only part of the variance (Engelhardt et al., 2015). The conservative nature of our models—such as controlling for fluid intelligence—likely attenuated the observed effects (Filippetti & Richaud, 2017). By excluding participants with cardiovascular disease—a group with higher trauma exposure and greater EF deficits—we may also have underestimated effect sizes. In addition, the middle-aged to older UKB cohort may show diminished adversity effects due to temporal decay or compensatory processes (Teicher et al., 2016). Beyond methodological considerations, small effect sizes at the individual level may accumulate to substantial functional impacts at the population level, particularly given the high prevalence of childhood adversity. Finally, volumetric measures may underestimate the true magnitude of adversity–EF associations, as functional or microstructural alterations (e.g., connectivity disruptions) could capture stronger effects.

This study has several limitations. First, the cross-sectional design prevents causal inference about the temporal sequence linking childhood adversity, brain structure, and EF. Second, reliance on retrospective trauma reports may introduce recall bias. Third, our assessment of EF was limited, as the UKB battery lacked inhibitory control and other core domains. Finally, the study focused on volumetric measures and did not account for functional connectivity or genetic influences, which may provide a more comprehensive understanding of adversity-related neural mechanisms.

Our findings demonstrate that distinct dimensions of childhood adversity leave separable neurocognitive signatures, providing mechanistic evidence that early environments sculpt executive systems in both maladaptive and adaptive ways, and establishing a framework for precision identification and intervention in the enduring cognitive sequelae of adversity.

## Data Availability

All data produced in the present work are contained in the manuscript

https://biobank.ndph.ox.ac.uk

